# Regional heterogeneity in the use of extracorporeal membrane oxygenation for heart transplant after 2018 allocation policy change

**DOI:** 10.1101/2025.02.07.25321902

**Authors:** Erin M. Schumer, Toyokazu Endo, Takuya Wada, Joel D. Schilling, Kunal D. Kotkar, M. Faraz Masood, Amit Pawale

## Abstract

**Background:** The 2018 heart transplant allocation policy has changed the profile of patients receiving heart transplant, with an increased use of acute mechanical support and decreased use of durable left ventricular assist devices. We investigated the use of extracorporeal membrane oxygenation (ECMO) use pre- and post-allocation change.

**Methods:** Using the UNOS database, we identified adult patients who underwent heart transplant from January, 2006 - June, 2022. The study time period was divided into pre- and post-allocation change. We divided ECMO patients by region and analyzed change in the rate of ECMO use by region between eras. Differences between groups and survival comparison were analyzed.

**Results:** A total of 41,636 recipients were found, of which 891 (2.1%) were on ECMO at the time of transplant. Overall ECMO use increased from 231 (0.8%) to 660 (5.5%) between eras (p<0.001). There was significant regional variation in the rate of ECMO usage (p<0.001) and significant differences in postoperative dialysis (p=0.014) and acute rejection episodes (p<0.001). There was no significant difference in pacemaker rate (p=0.172), stroke (p=0.212), treatment for rejection within 1 year (p=0.358), or post-transplant survival in the current era between regions for patients on ECMO at the time of transplant (p=0.444).

**Conclusion:** There is increased utilization of ECMO following implementation of an allocation system which prioritizes ECMO recipients on the waitlist and there <u>is a differential</u> increase in the use of ECMO in various regions. Further granular studies are needed to see which patients may benefit more from ECMO and what can be done to reduce post-transplant mortality from ECMO to transplant as it remains high before and after allocation change.

## Introduction

Healthcare policy affects patient outcomes, and the heart allocation policy is not an exception. In recent years, there has been controversy within the field of heart transplantation given the new heart allocation policy [1–4]. In 2018 Organ Procurement and Transplantation Network (OPTN) implemented a new heart allocation policy that introduced the six-tier system to reduce waitlist mortality and prioritize those that are the most vulnerable [5]. Various studies have looked at the outcomes of heart transplantation since the enactment of the new policy, and the current literature shows that the new policy has helped reduce the waitlist time and improve waitlist outcomes [2,6,7].

The new allocation policy split the previous model to give priority to those who are critically ill. As patients on temporary mechanical circulatory support (MCS) devices are given higher priority for transplant, the use of these devices has increased since the allocation system change. To <u>qualify</u> for status 1, the highest listing priority, requires the use of venoarterial extracorporeal membrane oxygenation (VA-ECMO), non dischargeable biventricular assist devices, or patients on MCS with arrhythmias. Stable patients on durable left ventricular assist devices (LVAD) were downgraded from previous status 1B to status 4, which has decreased the number of patients on LVADs receiving transplants [8]. These changes have dramatically shifted the distribution of patients with MCS at the time of transplant.

VA-ECMO is an invaluable tool to support critically ill patients with advanced heart failure with refractory cardiogenic shock [9] but is reserved for selected patients [10]. Since the implementation of the new allocation system, there has been an increase in utilization of VA-ECMO, and there has been an improvement in both waitlist time and waitlist mortality with similar post-transplant survival [11]. However, it is imperative to note despite the improvement, patients bridged to heart transplant with VA-ECMO tend to have worse outcomes compared to those without [12,13]. Thus, careful consideration must be employed to utilize VA-ECMO as a bridging strategy.

Since the policy change, there is evidence that there are geographical and regional variations in pre-transplant management [14]. Furthermore, Cascicino et al. have shown that there has been a significant increase in the utilization of temporary MCS in patients listed for heart transplantation with significant variance across different centers [15]. It is well known that geographical barriers exist regarding outcomes, which have been more prevalent since the new policy [16,17]. Thus, it is imperative to further evaluate the utility of VA-ECMO in patients listed for heart transplantation, as it has potential for both high risk of mortality and high reward to reduce wait times and even waitlist mortality. We examined regional patterns of ECMO use pre- and post-allocation change to further explore the impact of policy on heart transplant outcomes and regional variations of practice.

## Patients and Methods

This study was exempt from the Institutional Review Board as it used a de-identified, administrative database. Similarly, consent was not obtained. Using the United Network for Organ Sharing (UNOS) database, we identified adult patients (≥ 18 years) who underwent heart transplant with available data from January, 2006 - June, 2022. The study time period was divided into pre- (January 2006 - October 17, 2018, Era 1) and post- (October 18, 2018 - June, 2022, Era 2) allocation change and was stratified by UNOS region. We identified patients on ECMO support at the time of transplant to compare absolute numbers and the rate of change between eras. Demographic information for both donors and recipients was compared between regions for recipients on ECMO in Era 2 at the time of transplant. Finally, postoperative outcomes for recipients on ECMO at the time of transplant were compared between regions.

Continuous variables were assessed for normality using the Levene Test for equality of variances and presented as mean (standard deviation) for continuous parametric or median (interquartile range) for continuous non-parametric. Categorical variables are presented as N (%). The groups were then compared using the student’s *t*-test (continuous parametric) or Kruskal-Wallis test (continuous non-parametric) or the Chi-Square test (categorical). Linear regression was used to analyze changes in the rate of ECMO use by region between the two eras. Kaplan-Meier survival with log-rank analysis was used to compare the overall survival differences between groups. A p-value of 0.05 was considered significant.

## Results

Overall, ECMO use increased from 231 (0.8%) to 660 (5.5%) between eras (p<0.001), with a significant increase in ECMO utilization across all regions since the change in policy (**Figure 1).** The most significant increase between eras was observed in regions 1, 6, and 9 **(Figure 2)** while other regions, such as 3, 10, and 11, had the least significant difference between eras (p<0.001). Although each region significantly increased the use of ECMO, specific centers comprised most of the utilization within each region. For example, two centers in Region 2 had more ECMO utilization than the rest, and in Region 9, one center composed 47% of the region’s ECMO utilization compared to the other six transplant centers **(Figure 3).**

**Figure 1.**
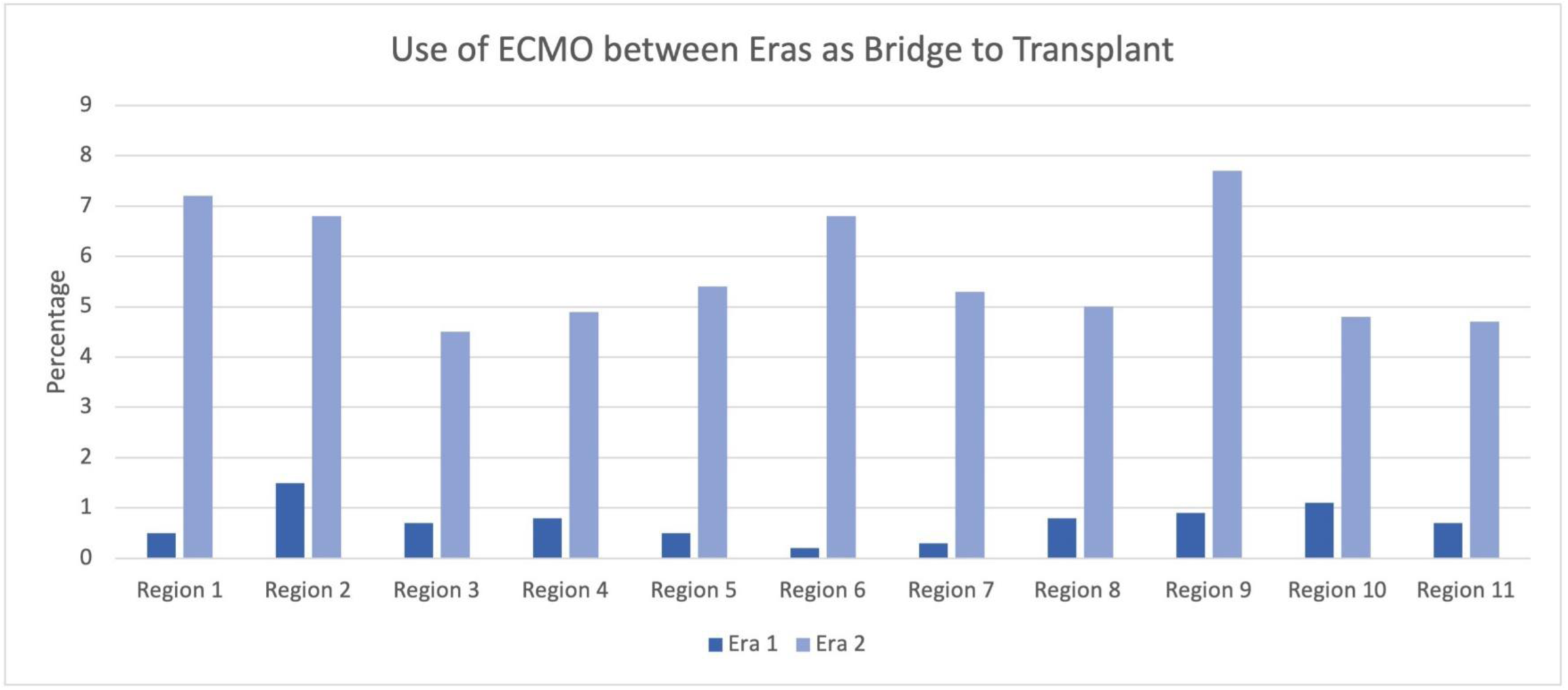
Histogram showing extracorporeal membrane oxygenation use by region pre and post 2018 allocation score change.

**Figure 2.**
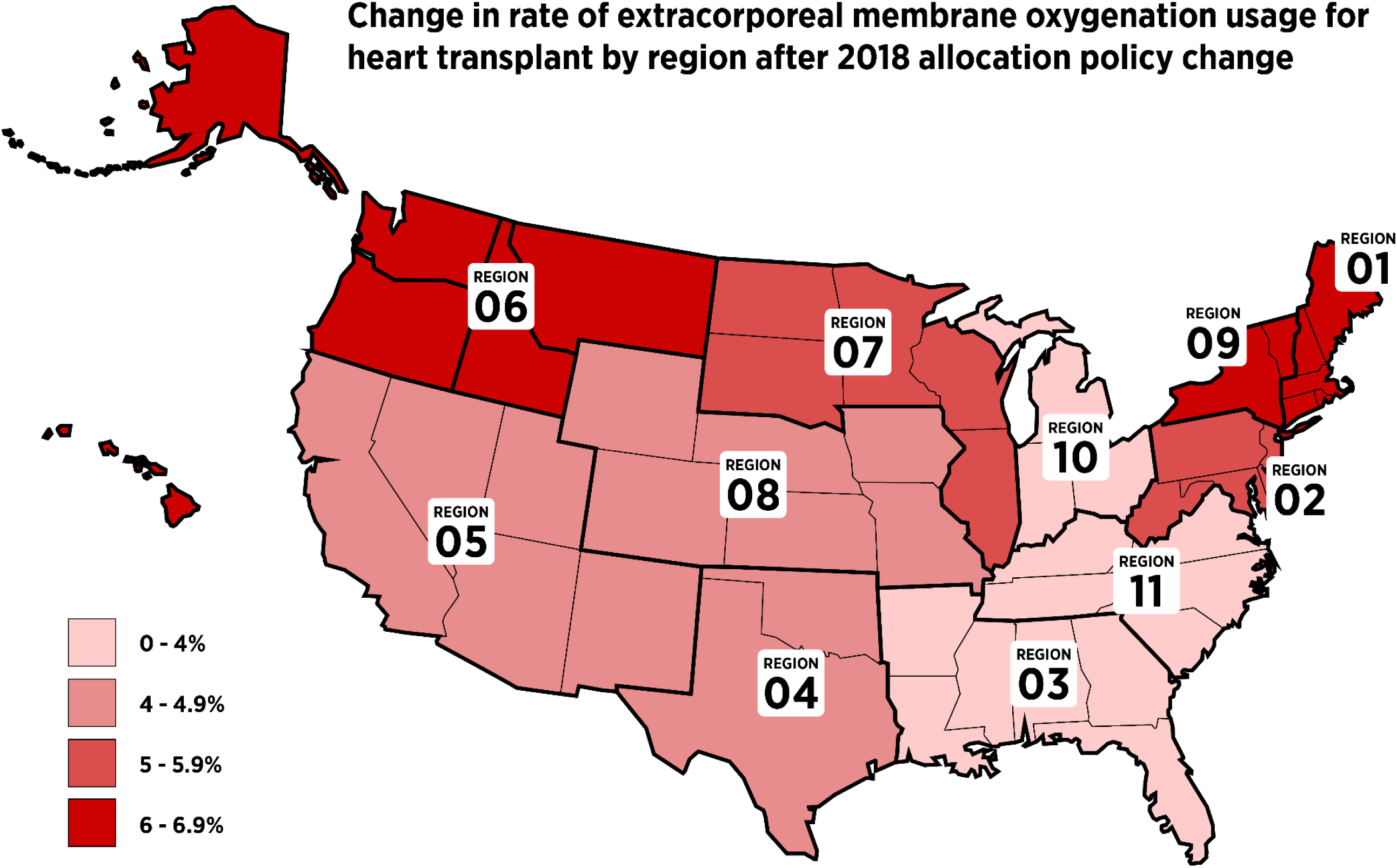
Change in the rate of extracorporeal membrane oxygenation from Era 1 (2006-October 18, 2018) to Era 2 (October 19, 2018 - June 30, 2022).

**Figure 3.**
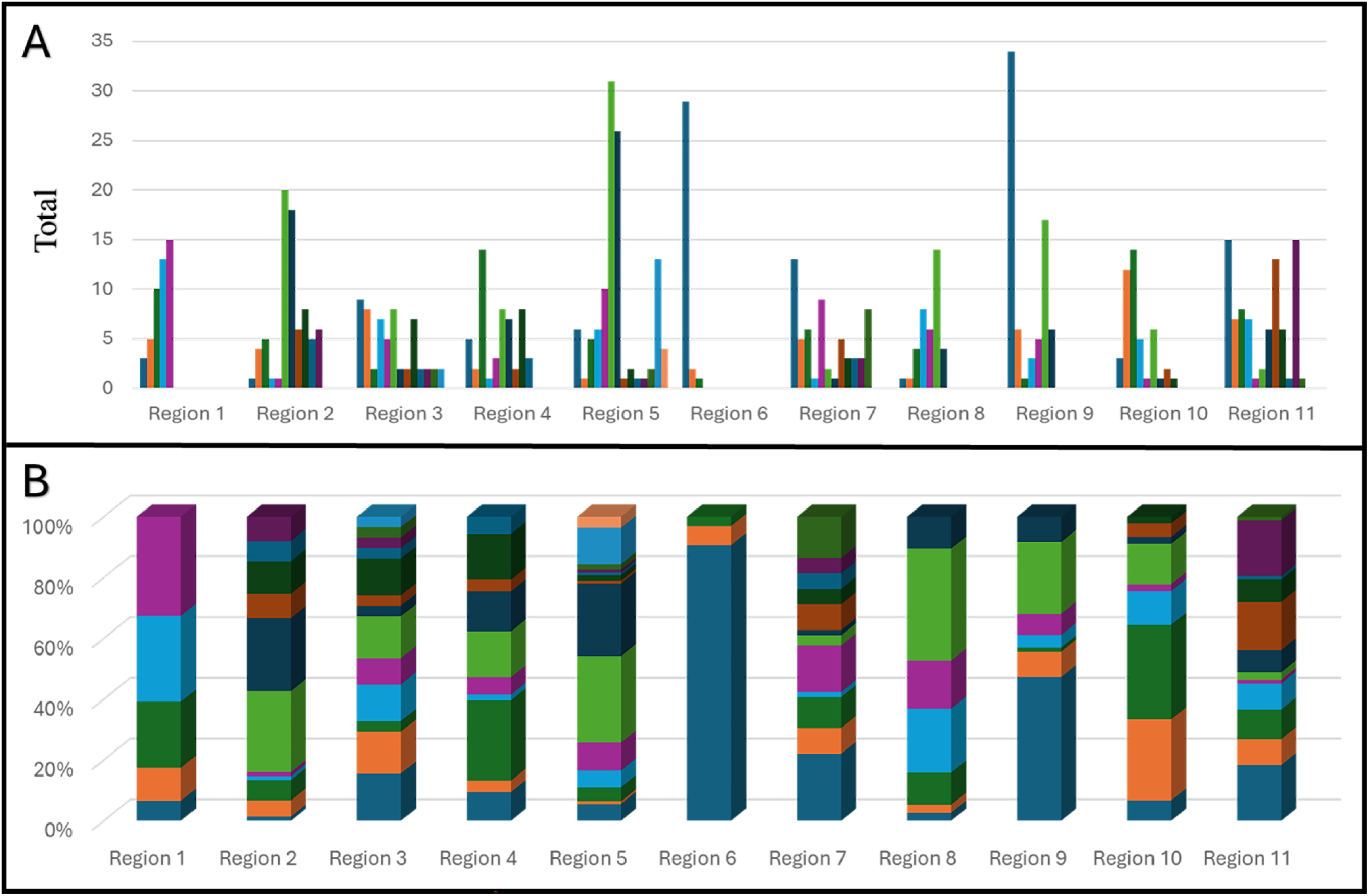
Regional extracorporeal membrane oxygenation (ECMO) use stratified by de-identified center. Figure A demonstrates the use of ECMO (frequency) per center per each UNOS region. The different colors indicate a different center within that region (Region 1 depicts five centers, while Region 11 depicts eleven centers). Figure B depicts the overall percentage of the total EMCO use per center in that region.

A total of 41,636 heart transplant recipients were performed, of which 891 (2.1%) were on ECMO at the time of transplant. Recipient characteristics for patients on ECMO support at the time of transplant are shown in **Table 1** and are similar between regions aside from ethnicity. Most recipients were white and male in all regions. Recipient history was also similar, with a similar proportion of diabetes (p=0.41), history of prior cigarette use (p=0.32), and preoperative dialysis use (p=0.44). The majority of recipients were listed for non-ischemic cardiomyopathy, with about a quarter of patients listed due to ischemic causes (p=0.64). There was no statistical difference in the median waitlist time across regions (p=0.59).

**Table 1.** Demographic characteristics for heart transplant recipients who were on extracorporeal membrane oxygenation at the time of transplant separated by United Network for Organ Sharing region. Results reported in N (%) or mean (± standard deviation) for parametric variables or median (interquartile range) for non-parametric variables. BMI = Body mass index, kg/m^2^ = kilogram per meter squared. * Denotes non-parametric testing

|  | Region<br>1 | Region<br>2 | Region<br>3 | Region<br>4 | Region<br>5 | Region<br>6 | Region<br>7 | Region<br>8 | Region<br>9 | Region<br>10 | Region<br>11 | p |
| --- | --- | --- | --- | --- | --- | --- | --- | --- | --- | --- | --- | --- |
| **Age (years)** | 48.4±14.2 | 44.5±15.3 | 45.4±14.8 | 51.1±13.0 | 48.3±14.5 | 48.6±15.5 | 46.9±17.3 | 44.5±14.3 | 47.6±15.2 | 48.2±14.9 | 46.2±14.5 | 0.138 |
| **Sex (% male)** | 42 (80.8) | 94 (71.8) | 66 (79.5) | 58 (73.4) | 99 (73.3) | 22 (88.0) | 41 (61.2) | 31 (71.2) | 68 (76.4) | 55 (77.5) | 79 (73.8) | 0.274 |
| **BMI (kg/m<sup>2</sup>)** | 26.9±5.2 | 26.1±5.0 | 27.5±5.9 | 27.2±4.4 | 26.2±5.0 | 27.4±5.9 | 28.3±5.3 | 27.7±5.1 | 27.1±5.1 | 27.3±5.1 | 27.1±5.7 | 0.229 |
| **Ethnicity**<br>White<br>Black<br>Other | 33 (63.5)<br>10 (19.2)<br>9 (17.3) | 86 (65.6)<br>33 (25.2)<br>12 (9.2) | 43 (51.8)<br>28 (33.7)<br>12 (14.5) | 45 (57.0)<br>16 (20.3)<br>18 (22.8) | 70 (51.9)<br>17 (12.6)<br>48 (35.6) | 20 (80.0)<br>3 (12.0)<br>2 (8.0) | 46 (68.7)<br>15 (22.4)<br>6 (9.0) | 41 (78.8)<br>9 (17.3)<br>2 (3.8) | 54 (60.7)<br>17 (19.1)<br>18 (20.2) | 60 (84.5)<br>9 (12.7)<br>2 (2.8) | 58 (54.2)<br>44 (41.1)<br>5 (4.7) | <0.001 |
| **Recipient history**<br>Diabetes<br>mellitus<br>Smoking history<br>Preoperative<br>dialysis | 10 (19.2)<br>12 (23.1)<br>4 (7.7) | 19 (14.6)<br>48 (36.6)<br>3 (2.3) | 22 (26.5)<br>26 (31.3)<br>12 (14.4) | 20 (25.4)<br>30 (38.0)<br>8 (10.2) | 32 (23.7)<br>43 (31.9)<br>16 (11.8) | 4 (16.0)<br>11 (44.0)<br>2 (8.0) | 17 (25.3)<br>21 (31.3)<br>11 (16.4) | 11 (21.1)<br>9 (17.3)<br>4 (7.7) | 19 (21.4)<br>22 (24.7)<br>9 (10.0) | 10 (14.1)<br>23 (32.4)<br>5 (7.0) | 34 (31.8)<br>32 (29.9)<br>9 (8.4) | 0.409<br>0.317<br>0.436 |
| **Creatinine**<br>**(mg/dL)** | 1.3±0.7 | 1.2±0.6 | 1.4±1.3 | 1.4±0.9 | 1.2±0.9 | 1.1±0.8 | 1.4±1.0 | 1.4±0.8 | 1.4±1.3 | 1.1±0.5 | 1.3±0.6 | 0.310 |
| **Time on waiting**<br>**list (days) \*** | 9 (3,43) | 9 (4,50) | 12 (3,39) | 8 (3,35) | 5 (3,12) | 5 (3,8) | 7 (4,31) | 4 (2,15) | 7 (3,25) | 7 (2,18) | 6 (3,16) | 0.587 |
| **Diagnosis**<br>Non-ischemic<br>Ischemic<br>Other | 29 (55.8)<br>12 (23.1)<br>11 (21.2) | 65 (49.6)<br>35 (26.7)<br>31 (23.7) | 43 (51.8)<br>15 (18.1)<br>25 (30.1) | 33 (41.8)<br>21 (26.6)<br>25 (31.6) | 70 (51.9)<br>27 (20.0)<br>38 (28.1) | 10 (40.0)<br>8 (32.0)<br>7 (28.0) | 30 (44.8)<br>15 (22.4)<br>22 (32.8) | 24 (46.2)<br>10 (19.2)<br>18 (34.6) | 43 (48.3)<br>24 (27.0)<br>22 (24.7) | 30 (42.3)<br>20 (28.2)<br>21 (29.6) | 60 (56.1)<br>28 (26.2)<br>19 (17.8) | 0.644 |

Donor characteristics are shown in **Table 2**. There was a significant difference in donor age (p=0.03) and no difference in the sex distribution (p=0.87). Most donors were white, with some regional variation (p<0.001). There was no statistical difference in BMI (p=0.22) or creatinine levels across regions (p=0.42). Donors’ history was also proportional across all regions. The cause of death was significantly different (p<0.001). Only regions 3, 9, and 11 had donors who underwent donation after circulatory death, but there was no statistical significance (p=0.48). In all regions, the donors’ median ejection fraction was similar (p=0.15). Ischemia time was significantly different (p=0.04), but the median ischemia time was < 4 hours for all regions.

**Table 2.** Postoperative outcomes for patients who were on extracorporeal membrane oxygenation at the time of heart transplant since the 2018 allocation policy change. ECMO= extracorporeal membrane oxygenation, PGF = primary graft failure. Treatment for rejection is for 1-year post-transplant. Reported in n (%)

|  | Region 1 | Region 2 | Region 3 | Region 4 | Region 5 | Region 6 | Region 7 | Region 8 | Region 9 | Region 10 | Region 11 | p |
| --- | --- | --- | --- | --- | --- | --- | --- | --- | --- | --- | --- | --- |
| Age (years) | 30.3±9.3 | 31.6±10.0 | 31.4±10.3 | 28.8±9.3 | 30.7±9.8 | 38.0±11.6 | 31.3±10.3 | 30.9±11.4 | 32.8±9.9 | 31.1±10.3 | 32.0±10.5 | 0.034 |
| Sex (% male) | 41 (78.8) | 98 (74.8) | 66 (79.5) | 61 (77.2) | 106 (78.5) | 21 (84.0) | 49 (73.1) | 39 (75.0) | 67 (75.4) | 55 (77.5) | 90 (84.1) | 0.867 |
| BMI (kg/m²) | 26.2<br>(23.1, 31.3) | 26.2<br>(22.6, 31.1) | 27.5<br>(24.3, 32.7) | 26.6<br>(23.8, 30.3) | 26.1<br>(23.9, 29.8) | 25.7<br>(23.7, 30.8) | 26.8<br>(23.7, 32.4) | 26.3<br>(23.8, 29.2) | 25.8<br>(22.9, 30.4) | 25.7<br>(22.9, 29.4) | 27.5<br>(24.1, 30.5) | 0.222 |
| Creatinine (mg/dL) | 1.0 (0.9, 2.2) | 1.1 (0.9, 1.8) | 1.0 (0.8, 1.8) | 1.0 (0.8, 1.5) | 1.2 (0.8, 1.8) | 1.0 (0.7, 2.3) | 1.0 (0.8, 1.5) | 1.1 (0.8, 1.4) | 1.0 (0.8, 1.5) | 1.0 (0.7, 1.3) | 1.0 (0.8, 1.5) | 0.419 |
| Ethnicity<br>White<br>Black<br>Other | 35 (67.3)<br>11 (21.2)<br>6 (11.5) | 90 (68.7)<br>23 (17.6)<br>18 (13.7) | 54 (65.1)<br>18 (21.7)<br>11 (13.3) | 43 (54.4)<br>13 (16.5)<br>23 (29.1) | 78 (57.8)<br>11 (8.1)<br>46 (34.1) | 13 (52.0)<br>3 (12.0)<br>9 (36.0) | 39 (58.2)<br>17 (35.4)<br>11 (1.4) | 37 (71.2)<br>8 (15.4)<br>7 (13.5) | 54 (60.7)<br>19 (21.3)<br>16 (18.0) | 48 (67.6)<br>16 (22.5)<br>7 (9.9) | 75 (70.1)<br>20 (18.7)<br>12 (11.2) | <0.001 |
| Donor History<br>Cocaine use<br>Smoking<br>Diabetes<br>Hypertension<br>Previous MI | 21 (40.4)<br>4 (7.7)<br>2 (3.8)<br>9 (17.3)<br>0 (0) | 31 (23.7)<br>14 (10.7)<br>5 (3.8)<br>16 (12.2)<br>0 (0) | 23 (27.7)<br>5 (6.0)<br>1 (1.2)<br>13 (15.7)<br>1 (1.2) | 13 (16.5)<br>7 (8.9)<br>1 (1.3)<br>12 (15.2)<br>2 (2.5) | 31 (23.0)<br>13 (9.6)<br>1 (1.4)<br>11 (8.1)<br>1 (0.7) | 4 (16.0)<br>6 (24.0)<br>1 (4.0)<br>5 (20.0)<br>2 (8.0) | 13 (19.4)<br>6 (9.0)<br>3 (4.5)<br>10 (14.9)<br>0 (0) | 13 (25.0)<br>4 (7.7)<br>0 (0)<br>5 (9.6)<br>0 (0) | 30 (33.7)<br>14 (15.7)<br>2 (2.2)<br>13 (14.6)<br>3 (3.4) | 17 (23.9)<br>12 (16.9)<br>0 (0)<br>8 (11.3)<br>0 (0) | 25 (23.4)<br>18 (16.8)<br>3 (2.8)<br>9 (8.4)<br>1 (0.9) | 0.254<br>0.264<br>0.504<br>0.856<br>0.062 |
| CDC High Risk | 21 (40.4) | 47 (35.9) | 22 (26.5) | 19 (24.1) | 33 (24.4) | 10 (40.0) | 18 (26.9) | 14 (26.9) | 34 (38.2) | 17 (23.9) | 23 (21.5) | 0.054 |
| Cause of Death<br>Anoxia<br>Cerebrovascular<br>Head trauma<br>CNS Tumor<br>Other | 29 (55.8)<br>3 (5.8)<br>19 (36.5)<br>1 (1.9)<br>0 (0) | 48 (36.6)<br>15 (11.5)<br>67 (51.1)<br>0 (0)<br>1 (0.8) | 38 (45.8)<br>8 (9.6)<br>36 (43.4)<br>0 (0)<br>1 (1.2) | 11 (13.9)<br>10 (12.7)<br>55 (69.6)<br>0 (0)<br>3 (3.8) | 48 (35.6)<br>21 (15.6)<br>58 (43.0)<br>0 (0)<br>8 (5.9) | 6 (24.0)<br>4 (16.0)<br>15 (60.0)<br>0 (0)<br>0 (0) | 26 (38.8)<br>10 (14.9)<br>29 (43.3)<br>0 (0)<br>2 (3.0) | 19 (36.5)<br>3 (5.8)<br>28 (53.8)<br>0 (0)<br>2 (3.8) | 43 (48.3)<br>11 (12.4)<br>34 (38.2)<br>0 (0)<br>1 (1.1) | 18 (25.4)<br>5 (7.0)<br>45 (63.4)<br>0 (0)<br>3 (4.2) | 37 (34.6)<br>13 (12.1)<br>55 (51.4)<br>1 (0.9)<br>1 (0.9) | <0.001 |
| Ejection Fraction | 60.5±6.5 | 60.9±6.4 | 62.5±6.3 | 60.1±6.7 | 62.3±7.0 | 63.3±5.3 | 60.9±5.0 | 62.1±6.5 | 62.0±6.6 | 62.1±7.4 | 61.2±5.6 | 0.147 |
| DCD Donor | 0 (0) | 0 (0) | 1 (1.2) | 0 (0) | 0 (0) | 0 (0) | 0 (0) | 0 (0) | 1 (1.1) | 0 (0) | 2 (1.9) | 0.480 |
| Ischemic Time (hours) | 3.4 (2.6, 3.9) | 3.3 (2.7, 3.7) | 3.5 (3.0, 4.3) | 3.4 (2.6, 3.9) | 3.6 (2.9, 4.1) | 3.5 (2.7, 5.0) | 3.4 (2.9, 4.0) | 3.4 (3.1, 3.9) | 3.2 (2.7, 3.6) | 3.5 (2.8, 3.9) | 3.5 (3.0, 3.8) | 0.044 |

There was a significant difference in postoperative use of dialysis among regions (p=0.014, **Table 3**). There was no significant difference in pacemaker rate (p=0.17) or stroke (p=0.21). All regions had less than five patients requiring pacemakers and less than 15 recipients who suffered from a stroke. There was a significant difference in rejection episodes with a disproportionate number of PGD, acute, and chronic rejections (p<0.001). However, there was no significant difference in the proportion of patients who received treatment for such rejections (p=0.36). There was no significant difference in post-transplant survival in the current era between regions for patients on ECMO at the time of heart transplant (p=0.444). However, when comparing the overall survival between era 1 and era 2, there was a significant improvement in region 3 (p=0.03), region 4 (p=0.001), region 9 (p=0.002), and region 11 (p=0.03, **Table 3**).

**Table 3.** Post-transplant overall survival differences within regions between eras for heart transplant recipients who were on extracorporeal membrane oxygenation at the time of transplant reported in mean days (95% confidence interval) and % survival.

|  | Era 1 |  | Era 2 |  | p |
| --- | --- | --- | --- | --- | --- |
| **Region 1** | 1795 (553, 3037) | 66.7% | 958 (835, 1081) | 87.0% | 0.206 |
| **Region 2** | 2713 (2099, 3327) | 51.8% | 852 (740, 965) | 79.7% | 0.256 |
| **Region 3** | 2350 (1454, 3247) | 44.0% | 826 (716, 937) | 81.0% | 0.025 |
| **Region 4** | 1320 (840, 1800) | 38.5% | 1026 (947, 1105) | 90.6% | 0.001 |
| **Region 5** | 3111 (1979, 4244) | 57.7% | 949 (859, 1038) | 82.4% | 0.760 |
| **Region 6** | 841 (0, 2007) | 50.0% | 975 (827, 1122) | 82.6% | 0.496 |
| **Region 7** | 1396 (182, 2610) | 37.5% | 923 (788, 1058) | 75.9% | 0.053 |
| **Region 8** | 3456 (2212, 4701) | 57.1% | 971 (859, 1083) | 86.1% | 0.543 |
| **Region 9** | 2427 (1427, 3427) | 52.9% | 1086 (1000, 1171) | 90.3% | 0.002 |
| **Region 10** | 3820 (2770, 4870) | 61.5% | 912 (787, 1037) | 82.2% | 0.283 |
| **Region 11** | 2519 (1499, 3538) | 44.0% | 861 (757, 965) | 79.3% | 0.029 |

## Comment

The 2018 heart allocation system change led to a drastically different “hemodynamic” profile of patients who receive a heart transplant in the current era. Since the allocation change, the use of temporary mechanical circulatory support has increased with fewer patients supported with durable LVADs. With the introduction of Impella 5.5 (Abiomed, Inc., Danvers, MA) in 2019, combined with the prioritization of the sickest patients for a heart transplant in the current allocation system, there has been a shift towards increased use of temporary mechanical support devices [18]. Although a small minority of patients, ECMO use has dramatically increased since the policy change. Thus we sought to describe regional differences in the use of ECMO pre- and post-allocation change.

Overall, our analysis showed a significant increase in the number of patients on ECMO at the time of transplant in the current allocation era, consistent with other reports [19–21]. However, the increase was not uniform between UNOS regions and appears to be driven by a few centers in each region. In the current allocation system, it is difficult to transplant patients on durable LVAD support, with wait times often exceeding two years [22,23]. Patients who were on ECMO in the previous era were likely to receive an LVAD as a bridge to transplant to optimize candidacy for eventual transplant [24]. With the change in prioritization, centers now have the option to directly transplant from ECMO; however, only the largest programs can support this from a programmatic standpoint as they may be more comfortable doing so given the large volume [25]. Recent reports have shown that adjusted outcomes are worse for patients on ECMO at the time of heart transplant as these patients are more likely to have an increased incidence of pulmonary hypertension, dialysis, stroke, and bleeding [26]. Our findings are consistent that overall, post-heart transplant survival is worse for patients who are on ECMO at the time of transplant compared with those who are not.

When examining post-transplant survival for only ECMO patients, there was no difference in survival between eras for most regions, but strikingly, post-transplant survival was improved in 4 of 11 regions. This improvement in survival is difficult to explain and still remains significantly worse than post-transplant survival for those not on ECMO at the time of transplant. A recent analysis of the Extracorporeal Life Support Organization (ELSO) database revealed that the acuity of patients cannulated for ECMO as a bridge to transplant has decreased since the 2018 allocation change [27]. Similarly, patients who had fewer complications related to ECMO were more likely to receive a transplant over an LVAD, which may be attributed to improvement in technology, increased options for left ventricular venting, limb perfusion, and anticoagulation management. What remains unexplained is why “less sick” patients are cannulated for ECMO in the current era compared with the pre-allocation change era. One explanation is that as more teams become comfortable with managing patients on ECMO, they are more likely to offer this therapy to patients in cardiogenic shock with the eventual hope of bridging to transplant or LVAD. Alternatively, some centers may favor ECMO support to allow patients to receive a transplant more quickly, especially if they have non groin cannulations, a program to mobilize ECMO patients and high level support from ECMO team. Some patients may also have needed ECMO support if deteriorated while awaiting transplantation on IABP or Impella support. Unfortunately, these explanations are speculative at this point and would require a deeper investigation into this phenomenon as time progresses under the new allocation system.

Our study has several limitations. The UNOS database is an administrative database. Thus, data is limited to the fields provided, and some data points are missing. We also do not have long-term follow-up for many patients as the allocation change went into effect in 2018; 5- and 10-year data will further elucidate the change in pre-operative support related to postoperative outcomes. Additionally, prior to the 2018 allocation change, very few patients were transplanted while on ECMO support, which makes robust conclusions challenging. Furthermore, we have demonstrated variation among UNOS regions, but it is imperative to state that the UNOS regions are not homogenous. Finally, the volume of data from the UNOS database helps to show overall trends. Still, the lack of intricate details related to this complex patient population makes it difficult to explain our study’s observations definitively.

In conclusion, we have shown significant regional variations in terms of ECMO use and bridging strategy. ECMO has provided a critical treatment option for those with severe heart failure, but it is not without significant risk. Though it makes sense to prioritize these patients on the waitlist, the center variation in ECMO use and patient characteristics raises the question of whether factors other than clinical need contributed to the higher use of ECMO as a bridging strategy. Heart transplants and the allocation of organs may not have been equitable in the current era. The new allocation system may need further revision to reduce geographical barriers and variations in managing those needing a heart transplant.

## Data Availability

The data are publicly available upon request from the OPTN database.

## Acknowledgments and Disclosures

The authors have no disclosures.

